# Financial barriers to accessing mental healthcare services among visually impaired people

**DOI:** 10.1101/2025.02.01.25321516

**Authors:** Annabelle A. Pan, Bani A. Aguirre, Bjorn K. Betzler, Mandeep S. Singh

## Abstract

**Aims:** Visual impairment (VI) is associated with heightened levels of loneliness, anxiety, and depression – yet few people with VI use mental healthcare services (MHS). The reasons for this are incompletely understood. Here, we aimed to understand possible financial barriers by analyzing the association of VI with delaying or foregoing MHS due to cost in a population database.

**Methods:** We conducted a cross-sectional study using the National Health Interview Survey (NHIS) of the U.S. Centers for Disease Control and Prevention (2019 to 2022). The primary outcome measure was the rate of delaying and foregoing obtaining MHS due to cost among participants with VI. Secondary outcomes included associations between sociodemographic characteristics and delaying or foregoing MHS due to cost. Logistic regressions were adjusted for demographic characteristics including household income.

**Results:** Among 18,475 participants, 27% reported VI. Participants with VI were more likely to be older, female, have lower income, and had lower educational attainment than those without VI. VI was associated with greater odds of delaying (adjusted odds ratio (aOR): 1.5, 95% confidence interval (CI) 1.3-1.6) and foregoing (aOR: 1.5, 95% CI 1.3-1.6) MHS due to cost. Among individuals with VI, those who were female and younger had higher odds of delaying and foregoing MHS due to cost.

**Conclusions:** Individuals with VI and depression or anxiety may be at higher risk of delaying or foregoing MHS due to cost. These findings underscore the importance of addressing financial barriers to ensure equitable access to MHS for individuals with VI.

## Introduction

The impact of vision impairment (VI) on mental wellbeing is widely appreciated. Previous work has shown that individuals with VI experience decreased health-related quality of life and are heightened levels of loneliness, anxiety, depression, psychological distress, and suicide.^1–7^ In tandem with the high prevalence of mood disorders, studies have reported an underutilization of mental healthcare services (MHS) among individuals with VI.^8^ In one study of 871 people with VI, a majority identified as needing MHS, but among those needing MHS only two thirds received any.^9^ A similar study found that although nearly 40% of 143 adults with VI screened positively for depressive symptoms, none currently received any kind of professional mental health support such as counseling.^10^ Another study involving 300 participants with age-related macular degeneration revealed that only one third of those with anxiety or depression symptoms reported receiving psychiatric treatment.^11^

These works underscore the large gap between the number of visually impaired people experiencing mental disorders and those who receive MHS as part of their medical care. Despite this gap, the factors contributing to the underutilization of MHS among people with VI are incompletely understood.

Several factors may contribute to the underutilization of MHS among people with VI. First, there generally exists a greater emphasis on physical rehabilitation following VI, with less attention given to emotional and psychological well-being.^12^ This prioritization of physical over mental rehabilitation may overshadow the mental health needs of individuals with VI. Additionally, many individuals with VI may lack awareness of how to access MHS, or may adopt a self-reliant approach to managing their emotional well-being, thus hindering help-seeking behaviors.^11,13^ Another critical barrier is the lack of expertise among MHS professionals in understanding of the unique challenges and experiences faced by individuals with VI, which may contribute to suboptimal care provision and reduced access to appropriate services.^14^

Our overarching goal was to understand why people with VI do not obtain MHS. It is known that people with VI experience more economic difficulty, including higher rates of living below the poverty line and experiencing food insecurity, compared to those without VI,^15^ yet the impact of cost as a barrier to accessing MHS among people with VI remains unexplored. Here, we aimed to study financial barriers as a possible reason for underutilizing MHS. This study leveraged population level data to assess the prevalence of cost barriers to mental healthcare services among individuals with VI.

## 2. Methods

### 2.1 Study Population

We analyzed publicly available and de-identified data for US adults (≥18 years) from the sample adult core questionnaire from the National Health Interview Survey (NHIS) administered by the United States CDC.^16^ The NHIS is an ongoing annual, cross-sectional, in-person and over-telephone household interview survey of the US noninstitutionalized civilian population. Due to a major redesign in 2019, only data from 2019 and onwards was included. The survey response rate ranged from 29.6% in the 2019-2020 longitudinal sample to 49.6% in 2022.^16^ Only individuals who reported a diagnosis of anxiety or depression were included to account for the confounding correlation between vision impairment and anxiety or depression.^5,6,17,18^

### 2.2 Ethics Statement

The study was not classified as human subjects research by the Johns Hopkins Institutional Review Board (IRB). It was exempt from IRB review because it analyzed existing publicly available data which was recorded in a way that participants cannot be identified. This study adhered to the guidelines of the Declaration of Helsinki. All respondents to NHIS provided oral consent before participation, and the survey was approved by the Research Ethics Review Board of the Centers for Disease Control and Prevention’s National Center for Health Statistics and the U.S. Office of Management and Budget.^16^

### 2.3 Exposure Definition

The main exposure was vision impairment (VI), defined as self-reported difficulty seeing. As done in previous studies, the presence of VI was defined as an affirmative response to the question: “Do you experience difficulty seeing, even when wearing glasses or contact lenses?”^1,15,19,20^ Respondents who answered affirmatively were categorized as having VI for the purposes of this study.

### 2.4 Outcome Definitions

The primary outcome assessed was the rate of facing cost barriers to mental healthcare services, defined as an affirmative response to: “During the past 12 months, have you delayed getting counselling or therapy from a mental health professional because of the cost?” or “During the past 12 months, was there any time when you needed counselling or therapy from a mental health professional, but did not get it because of the cost?”

Secondary outcomes assessed were the presence of mood disorder symptoms in participants, defined by individual responses to the questions: (1) “How often do you feel [worried, nervous, or anxious] OR [depressed]?” and (2) “Thinking about the last time you felt [worried, nervous, or anxious) OR [depressed], how would you describe the level of these feelings?”. As done in previous studies, respondents who answered: “daily” or “weekly” to the first question, and answered: “a lot” or “somewhere in between a little and a lot” to the second question, were categorized as having self-reported anxiety or depression symptoms.^21,22^

Other secondary outcomes were the usage of MHS, stratified by use of medication (defined as an affirmative response to: “Do you take prescription medication for (anxious feelings/ depression)?”) and therapy (defined as an affirmative response to: “During the past 12 months, did you receive counselling or therapy from a mental health professional such as a psychiatrist, psychologist, psychiatric nurse, or clinical social worker?”).

#### Other variables

Demographic variables included to adjust for confounding were sex, age, race, educational attainment, ratio of household income to poverty line (which is referred to as income ratio), and year of survey response. Health-related variables included having conditions of diabetes, hypertension, and heart disease.

### 2.5 Statistical analysis

We conducted a retrospective longitudinal analysis using survey-specific descriptive statistics to obtain weighted estimates for prevalence of mental healthcare symptoms, usage, and access, following the CDC’s NHIS-specific guidelines for pooling data across 2019 and onwards.^23^ Student t-tests and Chi square analyses were used to compare VI and non-VI populations. Multivariable logistic regression models were constructed using the PROC program in SAS to examine the association between VI and delaying or foregoing MHS, with adjustment for demographic variables, health-related variables, and mental health symptoms. No instances of collinearity were found. Missing data were excluded from analysis. All analyses were performed using SAS (SAS Institute, Inc., Cary, North Carolina). P-values were reported as 2-sided, and statistical significance was set at alpha = 0.05.

Subgroup analysis was performed by filtering the dataset to only include individuals with VI, then performing logistic regression between demographic variables and outcome measures within this dataset to identify populations that were more at risk of delaying or foregoing MHS due to the cost. Odds ratios were adjusted for all demographic variables that were not the variable of interest.

## Results

Of the 82,632 respondents to the combined 2019-2022 NHIS, 18,475 met inclusion criteria for having a self-reported diagnosis of anxiety or depression and a valid response to a VI question. Demographic characteristics of the study population are presented as weighted percentages in **Table 1**. Among all respondents, the raw and survey-weighted proportions of people with VI were 27.9% and 27.2% respectively. Compared to people without VI, people with VI were more likely to be female, older, and have lower levels of income and education. People with VI also had higher rates of diabetes, hypertension, and heart disease.

**Table 1.**
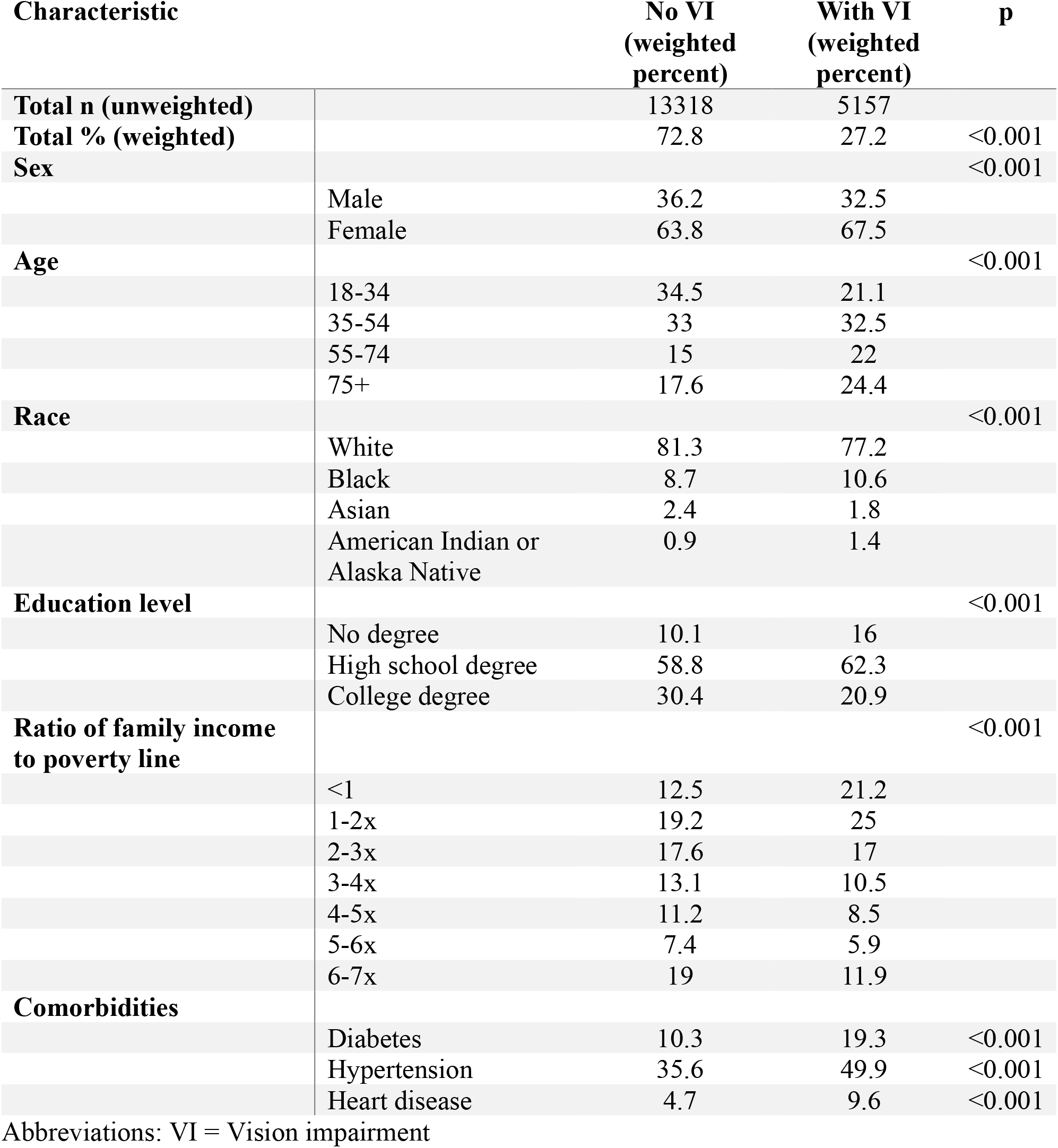
Demographics of study participants in weighted percentages.

The weighted prevalence of mood disorder symptoms, MHS usage, and delaying or foregoing MHS due to the cost are shown separated by people with and without VI in **Table 2**. After adjusting for demographic variables and health conditions, people with VI were more likely to delay (aOR: 1.5, 95% CI 1.3-1.6) and forego (aOR: 1.5, 95% CI 1.3-1.6) mental healthcare due to the cost compared to the non-VI group (**Table 3**). VI was associated with an increased frequency of anxiety or depression symptoms (aOR 1.5, 95% CI 1.4-1.6) (**Table 3**). People with VI were more likely to be taking medications for anxiety or depression (aOR: 0.9, 95% CI 0.9-0.998, p = 0.003). People with VI or without VI did not have significantly different rates for having seen a mental health provider in the past year (p = 0.84) (**Table 3**).

**Table 2.** Mental health characteristics of study participants.

| Characteristic |  | No VI<br>(weighted<br>percent) | With VI<br>(weighted<br>percent) | p |
| --- | --- | --- | --- | --- |
| **Self-reported diagnosis from**<br>**mental health professional** |  |  |  |  |
|  | Anxiety | 70.6 | 71.8 | 0.154 |
|  | Depression | 75.6 | 81.4 | <0.001 |
| **Mental health symptoms** |  |  |  |  |
|  | Either | 51.0 | 58.9 | <0.001 |
|  | Anxiety | 52.3 | 44.9 | <0.001 |
|  | Depression | 24.4 | 33.9 | <0.001 |
| **Taking medication for**<br>**depression or anxiety** | (yes) | 56.6 | 56.8 | 0.904 |
| **Received mental health**<br>**counseling in the past 12 months** | (yes) | 34.0 | 30.6 | <0.001 |
| **Has delayed counseling for**<br>**mental health due to cost in past**<br>**12 months** | (yes) | 12.1 | 14.0 | <0.001 |
| **Has foregone counseling for**<br>**mental health due to cost in past**<br>**12 months** | (yes) | 11.8 | 13.9 | <0.001 |

Abbreviations: VI = Vision impairment

**Table 3.** Association between vision impairment and mental health characteristics, before and after adjusting for mental health characteristics, demographic information, and comorbid chronic disease.

| **Mental health characteristic** | **OR (95% CI)** | **aOR (95% CI)** | **p (adjusted)** |
| --- | --- | --- | --- |
| Greater than weekly symptoms of anxiety or depression | 1.4 (1.3-1.5) | 1.5 (1.4-1.6) | <.001 |
| Taking medication for mental health | 1 (0.9-1.1) | 0.9 (0.9-1) | 0.003 |
| Received counseling in past 12 months | 0.9 (0.8-0.9) | 1 (0.9-1.1) | 0.836 |
| Delayed counseling/therapy due to cost | 1.2 (1.1-1.3) | 1.5 (1.3-1.6) | <.001 |
| Did not receive counseling/therapy due to cost | 1.2 (1.1-1.3) | 1.5 (1.3-1.6) | <0.001 |

Abbreviations: OR = odds ratio, aOR = adjusted odds ratio

In a subgroup analysis among only people with VI, female individuals had a significantly higher risk for delaying (aOR: 2.0, 95% CI 1.8-2.4) and foregoing (aOR: 2.1, 95%CI 1.9-2.5) MHS compared to male individuals (**Table 4**). Asian individuals had the lowest risk, followed by Native American individuals, Black individuals, and White individuals having incrementally higher risk for foregoing mental healthcare due to the cost (**Table 4**). Those aged 65+ were the least likely to report foregoing mental healthcare due to cost (aOR: 0.1 95% CI 0.1-0.2) compared to younger individuals aged 18-34 (**Table 4**). Individuals with older age reported lower likelihood of delaying or foregoing cost in a dose-dependent manner (**Table 4**). The likelihood of foregoing MHS due to the cost was associated with lower family income and higher education level (**Table 4**). The year of survey administration and comorbid chronic illnesses did not correlate strongly with delaying or foregoing care among those with VI (**Table 4**).

**Table 4.**
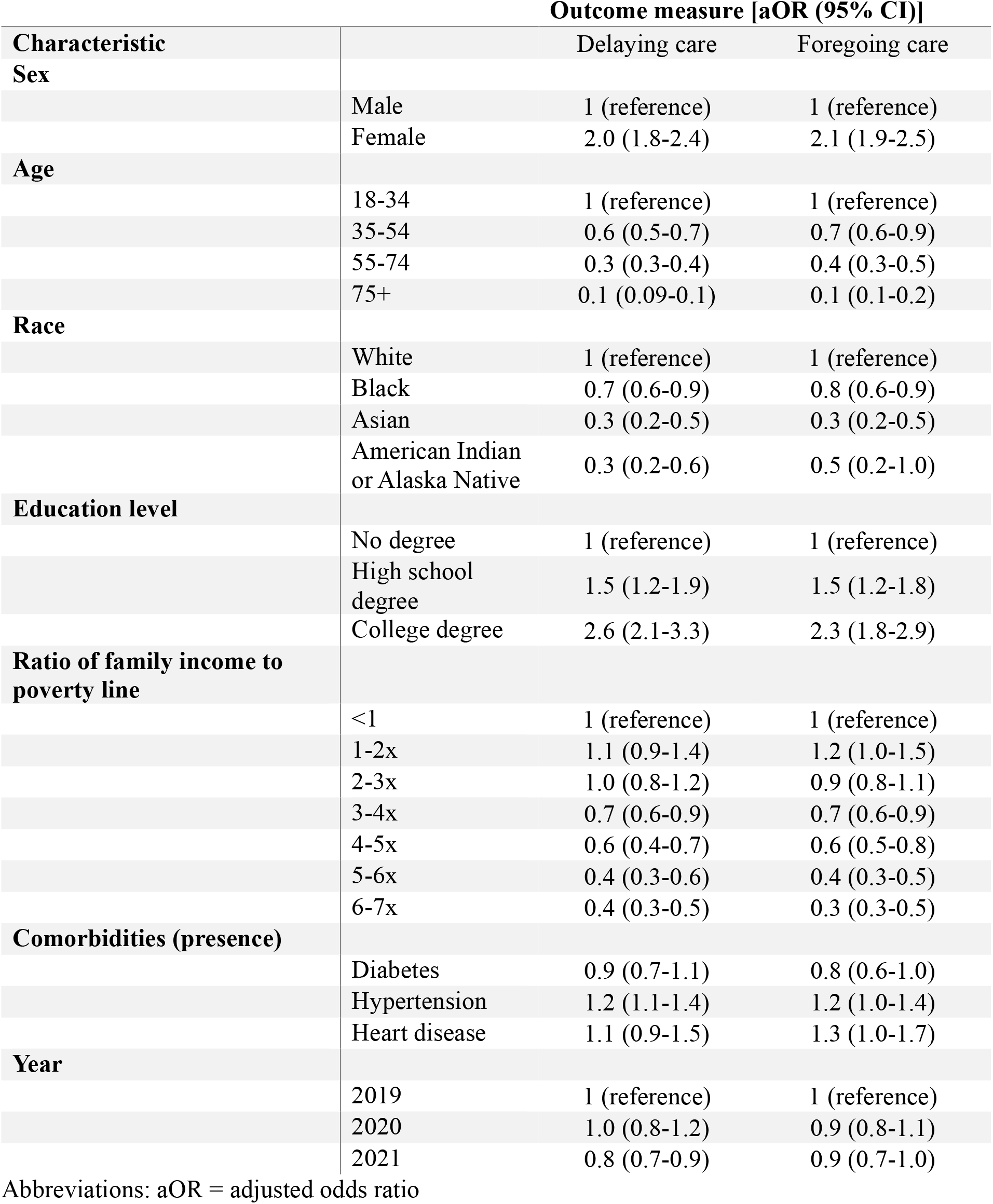
Association between demographic characteristics and delaying or foregoing mental healthcare services among people with vision impairment, adjusting for mental health characteristics, demographic information, and comorbid chronic disease.

## Discussion

This study investigated the association between self-reported VI and facing financial barriers to mental healthcare services. We found that individuals with VI reported increased rates of delaying or foregoing mental health services due to financial constraints, both before and after controlling for demographic factors including household income and comorbid chronic illnesses. The findings indicate that there may be substantial cost-related barriers to accessing mental healthcare services among people with VI and concomitant mood disorder.

To our knowledge, data on cost barriers to MHS utilization among individuals with VI have not been published. Other studies have reported the under-utilization of MHS among individuals with VI to be associated with a perceived de-emphasis on mental health in favor of visual health, a tendency towards self-reliance rather than help-seeking among people with VI, and a limited supply of mental health providers with experience and willingness to treat individuals with low vision.^11–14^ Our study uncovers and highlights the cost of mental healthcare as a significant contributor to the complex and possibly interconnected reasons why people with VI may not get the mental healthcare they need.

Additionally, we found that individuals with VI had a comparable level of MHS usage compared to those without VI, as represented through the frequency of seeing a mental health professional or taking prescription medication for a mental health condition. However, people with VI also reported higher rates of anxiety and depression symptoms compared to people without VI. This underscores the possible under-utilization of MHS among individuals with VI, who may have similar levels of use but a greater level of need for MHS than people without VI. These findings are consistent with previous studies that identified non-financial barriers to accessing mental healthcare access among people with VI.^9–11^

In our subgroup analysis of individuals with VI, age emerged as a major factor associated with the experience of cost-related barriers to MHS. Younger individuals with VI reported higher rates of delaying or foregoing mental health services due to financial limitations. This finding might be linked to an increased experience of mental health symptoms among younger individuals with VI.^1^ Interestingly, individuals who are White and have higher levels of education are more likely to report delaying and foregoing MHS due to cost. This could reflect a difference in individual perceptions of mental healthcare, which is generally more accepted in white and educated settings. Other settings may emphasize a reliance on oneself and one’s community for mental health, and previous work has found that non-White individuals with VI report lower levels of having satisfactory social support. This could indicate a difference in perception towards professionally-provided mental health services, rather than a difference in the overall mental health of these populations.

The present study has several limitations. First, the reliance on self-reported variables to represent vision impairment and mood disorder symptoms introduces an inherent subjectivity, as self-reported variables are imprecise analogs to clinical diagnoses. Nevertheless, there is strong concordance between self-reported and clinically-diagnosed vision impairment and moderate concordance for mood disorders, indicating general validity of relying on self-reported variables for research purposes.^24–27^ However, the intrinsic vagueness of the phrase, “difficulty seeing,” is a limitation inherent of the study, which cannot be uniformly generalized to the myriad causes of vision impairment. Additionally, with regards to the assignment of sex among participants, the NHIS provides only response options of “male,” “female,” and “don’t know,” which prevents the documentation of accurate information regarding gender identify for trans and nonbinary individuals. Future research should integrate more clinically precise variables including verified visual function data for each individual. Additionally, the population sampled includes only non-incarcerated adults in the United States, and conclusions may not be generalized beyond this population.

Identifying perceived financial constraints as a key obstacle to obtaining sufficient mental healthcare suggests a potential for informing patients and subsidizing care or assistance programs to improve accessibility for individuals with VI. As research continues to evolve in this area, future studies should incorporate more refined variables, enabling a more nuanced understanding of the relative impact of various barriers to MHS utilization among the diverse population of individuals with VI.

## Conclusions

Individuals with VI and concomitant depression or anxiety may be at higher risk of delaying or foregoing MHS due to cost-related reasons. The gap between MHS need versus its utilization among people with VI suggests a need to mitigate cost and other healthcare access barriers faced by persons living with VI and mental illness. Our findings underscore the importance of addressing financial barriers to accessing comprehensive care that includes MHS, to ensure equitable access to healthcare services among individuals with VI living with mental health challenges.

## Data Availability

All data produced in the present work are contained in the manuscript

## Author contributions

AAP and MSS conceived the study; AAP, BAA, BKB acquired and/or analyzed data; AAP, BAA, BKB drafted the initial manuscript; all authors provided critical review and approved the final manuscript.

